# Galectin approach to lower covid transmission - Drug Development for clinical use

**DOI:** 10.1101/2022.11.09.22282151

**Authors:** Alben Sigamani, Kevin H Mayo, Hana Chen-Walden, Surender Reddy, David Platt

## Abstract

**Background:** SARS-CoV-2 vaccines play an important role in reducing disease severity, hospitalization, and death, although they failed to prevent the transmission of SARS-CoV-2 variants. Therefore, an effective inhibitor of galectin-3 (Gal-3) could be used to treat and prevent transmission of COVID-19. ProLectin-M (PL-M), a Gal-3 antagonist, has been shown to interact with Gal-3 and thus prevent cellular entry of SARS-CoV-2 in previous studies.

**Aim:** The present study aimed to further evaluate the therapeutic effect of PL-M tablets in 34 subjects with COVID-19 disease, in addition to determining the mechanism of PL-M in preventing SARS-CoV-2 cell entry by NMR studies.

**Methods:** The efficacy of PL-M was evaluated in a randomized, double-blind, placebo-controlled clinical study in patients with mild to moderately severe COVID-19. Primary endpoints included changes in absolute RT-PCR Ct values of the nucleocapsid and open reading frame (ORF) genes from baseline to days 3 and 7. The incidence of adverse events, changes in blood biochemistry, inflammatory biomarkers, and levels of antibodies against COVID-19 were also evaluated as part of the safety evaluation. *In vitro* ^1^H-^15^N HSQC NMR spectroscopy studies were also performed to determine the interactions of PL-M with Gal-3 and the S1 spike protein of SARS-CoV-2.

**Results:** PL-M treatment significantly (p = 0.001) increased RT-PCR cycle counts for N and ORF genes on days 3 (Ct values 32.09 and 30.69 ± 3.38, respectively) and 7 (Ct values 34.91 ± 0.39 and 34.85 ± 0.61, respectively) compared to placebo. On day 3, 14 subjects in the PL-M group had cycle counts for the N gene above the cut-off of 29 (target cycle count 29), while on day 7 all subjects had cycle counts above the cut-off. Ct values in placebo subjects were consistently less than 29, and no placebo subjects were RT-PCR negative, until day 7. ^1^H-^15^N HSQC NMR spectroscopy revealed that PL-M specifically binds Gal-3 in the same way as the structurally similar NTD of the SARS-CoV-2 S1 subunit.

**Conclusion:** PL-M is safe and effective for clinical use in reducing viral load and promoting rapid viral clearance in COVID-19 patients by inhibiting SARS-CoV-2 entry into cells through inhibition of Gal-3.

## 1. Introduction

The COVID-19 disease, caused by the severe acute respiratory syndrome coronavirus 2 (SARS-CoV-2), remains an unprecedented event in world history. More than 2 years after the COVID-19 outbreak, the number of infections continues to rise in many areas of the world. Despite the availability of vaccines and other approved treatments, the prolonged persistence of the COVOD-19 disease is a major concern for the economy and people’s quality of life. However, existing vaccines continue to be extremely effective in reducing disease severity and preventing hospitalization and death [1]. The continued emergence of highly transmissible or pathogenic multiple SARS-CoV-2 variants with the potential to evade existing COVID-19 vaccines and antibody therapies contributes to disease transmission and persistence [2,3]. Potent antiviral regimens that rapidly reduce SARS-CoV-2 viral load and accelerate viral clearance in patients may hold promise, with the goal of preventing viral transmission and the emergence of new variants. In this context, a novel molecular approach that targets viral entry by engaging galectins is expected to be promising.

Galectins are small S-type lectins that function as pattern recognition receptors to enhance microbial invasion and regulate innate immune responses [4,5]. These selectively bind N-acetyllactosamine (Gal1-3GlcNAc or Gal1-4GlcNAc) of N-linked and O-linked glycoproteins [6,7]. Among the 12 human galectins, Galectin-3 (Gal-3) is expressed abundantly during viral infections in various immune cells and tissues and regulates their entry and attachment [8], replication [9], and inflammatory responses [10]. Recently, the role of Gal-3 in SARS-CoV-2 disease severity and associated cytokine storm syndrome has been described [11,12]. Recognition and binding to cell receptors is a critical step in viral infection. The S1 subunit of the SARS-CoV-2 spike protein consists of an N-terminal domain (NTD) and a C-terminal domain (CTD). Both facilitate viral adherence and entry processes [13] by binding with host ACE2 receptors in a S1-CTD fashion [14], while S1-NTD binds with N-acetylneuraminic acid (Neu5Ac) sugar receptors, Gal-3, and sialic acid-linked GM1 ganglioside [15]. The galactose-binding domain of human Gal-3 showed a nearly identical topology with the S1-NTD of SARS-CoV2 (Li, 2015) and also showed higher binding affinity with GM1 ganglioside [16]. It has also been reported that both Neu5Ac and the carbohydrate recognition domain (CRD) of Gal-3 receptors have a COOH terminal which strongly interacts with sugar molecules such as lactose and larger galacto-oligosaccharides or protein molecules including SARS-CoV2 spike protein [15,17]. The abundance of Neu5Ac and GAL-3 in the human nasopharynx and oral mucosa [18], and their strong interaction with protein molecules, may lead to high transmissibility and infectivity of SARS-CoV-2, particularly at viral entry points [19, 20]. Thus, understanding the numerous roles of Gal-3 in viral infection, it is believed that human Gal-3 antagonists are capable of preventing SARS-CoV2 adhesion and cell entry, as well as virus-associated inflammatory responses.

The RT-PCR technique is performed to determine the viral loads in infected patients that correspond to PCR cycle thresholds (Ct values). Though it may not be relevant for clinical outcomes, it is an important measure of the SARS-CoV-2 infectivity of an individual from the threshold cycle (Ct) value [21]. Hence, any intervention shown to increase the cycle threshold, indicating a reduction in viral multiplication and thus infectivity, is significant. ProLectin-M (PL-M) is a novel polymeric carbohydrate derived from gaur gum that competitively binds to the NH2 terminal domain (NTD) of human Gal-3 [22]. In our previous pilot study on 10 patients with COVID-19 infection, we found that oral administration of PL-M (chewable tablets) increased PCR cycle threshold, indicating the elimination of SARS-CoV-2 as well as a reduction in viral load in patients [23]. Further in vitro studies on SARS-CoV-2-infected Vero cells, followed by NMR analysis, revealed that PL-M strongly binds to human Gal-3, resulting in a reduction in viral load in Vero cells [24]. In this study, we further share the results of a clinical trial on CODID-19 patients and an NMR experiment with the extracted spike proteins.

## 2. Methods and Materials

### 2.1. Study design and oversight

The efficacy of the study drug (Chewable Tablet Galactomannan, 1,400 mg PL-M) was assessed in a randomised, double-blind, placebo-controlled clinical trial in ambulatory patients with mild to moderately severe COVID-19 infection. The study protocol was approved by the Institutional Ethics Committee (IEC) of ESIS Medical College and Hospital, Sanath Nagar, Hyderabad, India (ESIC Registration No: ECR/1303/Inst/TG/2019). As per regulatory standards, the study was registered with the Clinical Trials Registry-India (CTRI/2022/03/040757) on March 3, 2022. The study was conducted in accordance with the Declaration of Helsinki, the Good Clinical Practice guidelines, and local regulatory requirements. Prior to any trial-related activity, written informed permission from each participant was obtained. A total of 34 participants were identified, screened, and enrolled in this study. A paper-based case report form was used to capture all the clinical data.

### 2.2. Inclusion and exclusion criteria

The study involved 34 participants aged ≥18 years with mild to moderately severe COVID-19 infection who were willing to provide written informed permission and follow the trial guidelines. Participants were enrolled in the study if they had a recent rRT-PCR positive diagnosis (≤ 3 days) for COVID-19 infection with any of the following conditions: Ct value ≤ 25, hospitalisation for classical (CDC defined) COVID-19 symptoms (onset ≤ 5 days), and high-risk category of morbidity with SARS-CoV-2 infection. The exclusion criteria for participation included oxygen saturation levels (SpO2) ≤ 94% on room air, pregnant or breastfeeding women, active malignancy or chemotherapy, known allergies to any component of the study intervention, and pre-existing medical conditions that made the study protocol unsafe to follow. Participants who were receiving or had received any investigational COVID-19 treatment within 30 days before screening were also excluded from this study.

### 2.3. Study objective and outcome measures

The objective of this study was to determine the efficacy of PL-M chewable tablets (1400 mg) as a galectin antagonist in patients with mild to moderately severe COVID-19 infection. The primary endpoint was to assess the change in the absolute counts of the nucleocapsid gene, open reading frame (ORF) gene and an increase in Ct value, estimated from COVID-19 samples taken from a nasopharyngeal swab. In addition, we intended to assess the cumulative incidence of adverse events (AEs), changes in clinical biochemistry, clinical haematology, changes in blood markers of inflammation and changes in COVID-19 antibody levels.

### 2.4. Randomization and treatment

Patients who signed the informed consent form and met the eligibility criteria were enrolled in the study. Enrolled participants were randomly assigned to either the PL-M or placebo groups in a 1:1 ratio. An independent biostatistician provided a computer-generated assignment randomization list and blocks with varied block sizes to the investigators. Patients were given 1 tablet (either PL-M or matching placebo, 1400 mg) every hour for 7 days, with a maximum dose of 10 tablets per day because the viral replication cycle is 8-10 hours. Each participant was instructed to hold the tablet in their mouth for 1-2 minutes before dissolving and swallowing it. During mealtimes, such as breakfast, lunch, tea, and dinner, the subject was required to wait 30 minutes after the previous meal before taking the next tablet. This was done to avoid any potential reduction in blood glucose levels caused by the tablets’ ability to impede absorption of carbohydrates taken during the meal.

### 2.5. Study interventions

The study interventions, PL-M (derived from gaur gum galactomannan) and placebo tablets (chewable), were manufactured at the GMP facility of Murli Krishna Pharma Pvt. Ltd., in Pune, Maharashtra. All the study interventions were identical in appearance, shape, colour, packaging, and texture.

### 2.6. Study procedures

At all visits on days 1, 3, and 7, nasopharyngeal/oropharyngeal swab samples were taken from each patient to test for COVID-19 positivity using the RT-PCR method. Swab samples were transported to the research facility for Ct value analysis for the ORF and N protein genes. Throughout the analysis, all laboratory workers remained blind to treatment allocation. RNA extractions were performed using the QIAamp Viral RNA mini kit (#52904, Qiagen) according to the manufacturer’s standard protocol. A sample was considered negative if no Ct value was obtained and no amplification curve was observed, or if the Ct value for all three targets was > 29. At each visit, clinical symptoms, adverse events (AEs), and concomitant medicines were recorded. Patient safety was assessed both at the beginning and completion of the trial (day 7). Changes in vital signs and laboratory tests such as haematology, and serum biochemistry were evaluated as part of the safety assessment. Patients were monitored for 28 days from the day of randomization.

#### 2.6.1. rRT PCR

The extracted RNA was analysed using TRUPCR® SARS-CoV-2 RT qPCR KIT V-2 (#3B3043B Black Bio Biotech). In the real-time RT-PCR assay, 2 target genes including ORF and N genes were evaluated. Ct values obtained from a series of 5 template DNA dilutions of at least 3 separate samples were graphed on the y-axis versus the log of the dilution on the x-axis to measure the efficiency of the PCR. The Ct values assumed by the following equation were used to determine the logarithm of the recombinant gene copy numbers from: Ct = slope log (Gene Copy Number) + 1, where 1 act as the standard curve’s intercept.

### 2.7. NMR spectroscopy

NMR studies were carried out at 30°C on a Bruker 850 MHz spectrometer outfitted with a H/C/N triple-resonance probe and an x/y/z triple-axis pulse field gradient unit. A gradient sensitivity-enhanced version of two-dimensional ^1^H-^15^N HSQC was applied with 256 (t1) x 2048 (t2) complex data points in nitrogen and proton dimensions, respectively. Raw data were converted and processed by using NMRPipe and were analyzed by using NMRview.

Uniformly 15N-labeled galectin-3 (Gal-3) was dissolved at a concentration of 20 µM in a 20 mM potassium phosphate buffer at pH 6.9, prepared with a 95% H2O/5% D2O mixture. To explore the binding of PL-M and its separate polysaccharide components (galactomannans AG and R) to Gal-3, ^1^H–^15^N HSQC NMR measurements were performed. The 1H and 15N resonance assignments of recombinant human Gal-3 were reported earlier.

### 2.8. Statistical analysis

A total of 34 subjects were randomized, and no formal sample estimation was undertaken. Ct values and absolute copy numbers were compared using a parametric, unpaired repeated t-test with Welch’s correction or a non-parametric Mann Whitney U test. A two-tailed, p<0.05, was considered statistically significant.

## 3. Results

### Baseline characteristics

Thirty-four participants with mild to severe COVID-19 disease were screened for this study, and all met the inclusion criteria. Table 1 summarizes the baseline characteristics of participants. Following randomization, each group had 17 participants, and all the participants had completed the study. The participants were 70.59% (24) male and 29.41% (10) female. The average age of the participants in the control group was 37.29 ± 7.73 years, while in the PL-M group it was 41.82 ± 5.27 years. There was no evidence of co-morbidity in any of the participants.

**Table 1:**
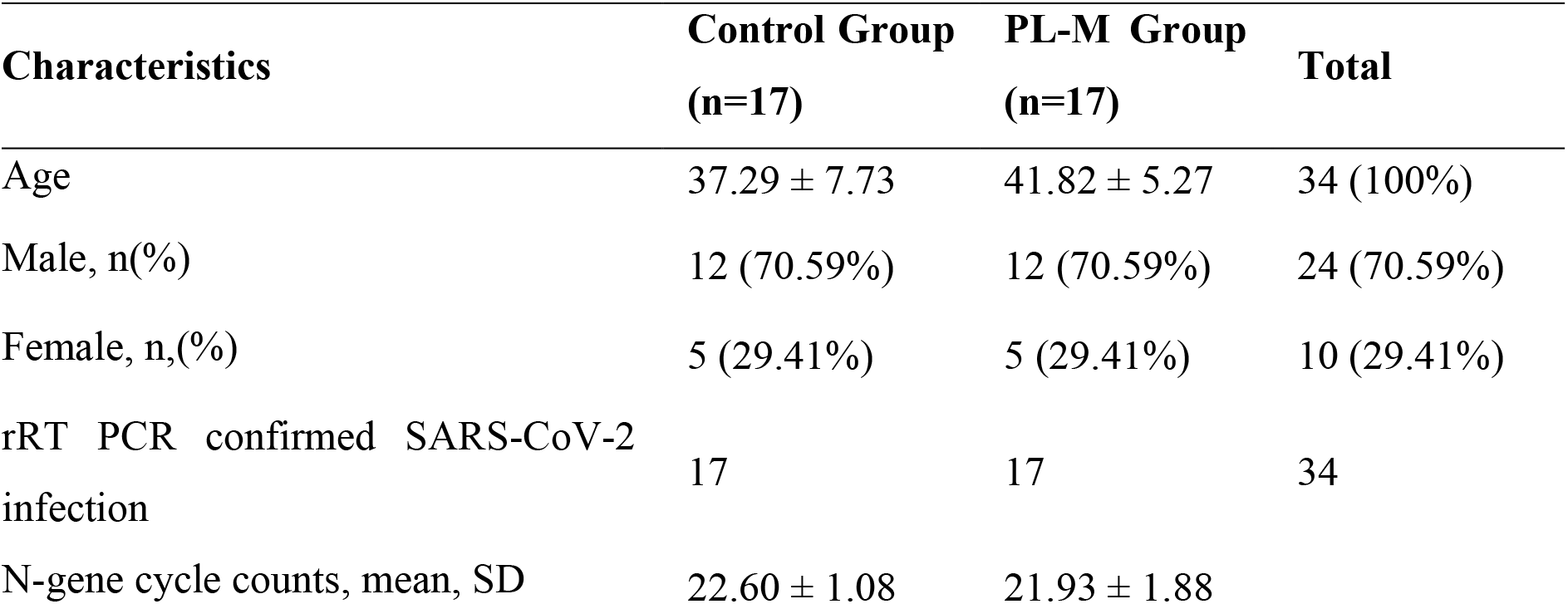
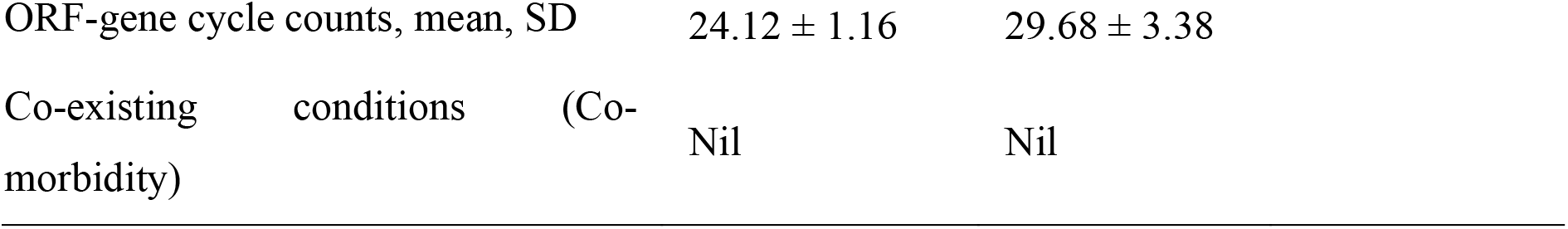
Baseline data

| Characteristics | Control Group<br>(n=17) | PL-M Group<br>(n=17) | Total |
| --- | --- | --- | --- |
| Age | $37.29 \pm 7.73$ | $41.82 \pm 5.27$ | 34 (100%) |
| Male, n(%) | 12 (70.59%) | 12 (70.59%) | 24 (70.59%) |
| Female, n,(%) | 5 (29.41%) | 5 (29.41%) | 10 (29.41%) |
| rRT PCR confirmed SARS-CoV-2 infection | 17 | 17 | 34 |
| N-gene cycle counts, mean, SD | $22.60 \pm 1.08$ | $21.93 \pm 1.88$ | |

|  |  |  |
| --- | --- | --- |
| ORF-gene cycle counts, mean, SD | 24.12 ± 1.16 | 29.68 ± 3.38 |
| Co-existing conditions (Co-morbidity) | Nil | Nil |

### Assessment of efficacy

SARS-CoV-2 RT-PCR tests were performed on days 1, 3, and 7. The results of the RT-PCR assays are presented in Table 2 and Figure 1A-C. As demonstrated in Table 2 and Figure 1A, B, the RT-PCR cycle counts for N and ORF genes in both groups increased during the treatment period (days 1-7). Mean RT-PCR cycle counts for both N and ORF genes were significantly (p = 0.001) higher in the PL-M treatment group in days 3 (Ct values 32.09 ± 2.39 and 30.69 ± 3.38, respectively) and 7 (Ct values 34.91 ± 0.39 and 34.85 ± 0.61, respectively). Contrarily, during the course of the study, mean RT-PCR cycle counts in the placebo group were consistently below 29 (targeted cycle count 29) for both N and ORF genes in days 3 (Ct values 24.38±1.25 and 24.12±1.16, respectively) and 7 (Ct values 27.24 ± 1.25 and 26.56 ± 1.22, respectively). On day 3, as can be seen from Figure 1C, RT-PCR cycle counts were above the targeted cut-off value of 29 in 14 or 82.35% of the participants in the PL-M treatment group, whereas all the participants in the placebo group had RT-PCR cycle counts below the targeted cut-off value. Interestingly, on day 7, all the participants in the PL-M treatment group had cycle counts above the targeted cut-off value, while the majority of participants (16, or 94.12%) in the placebo group had cycle counts below the targeted cut-off value.

**Table 2:**
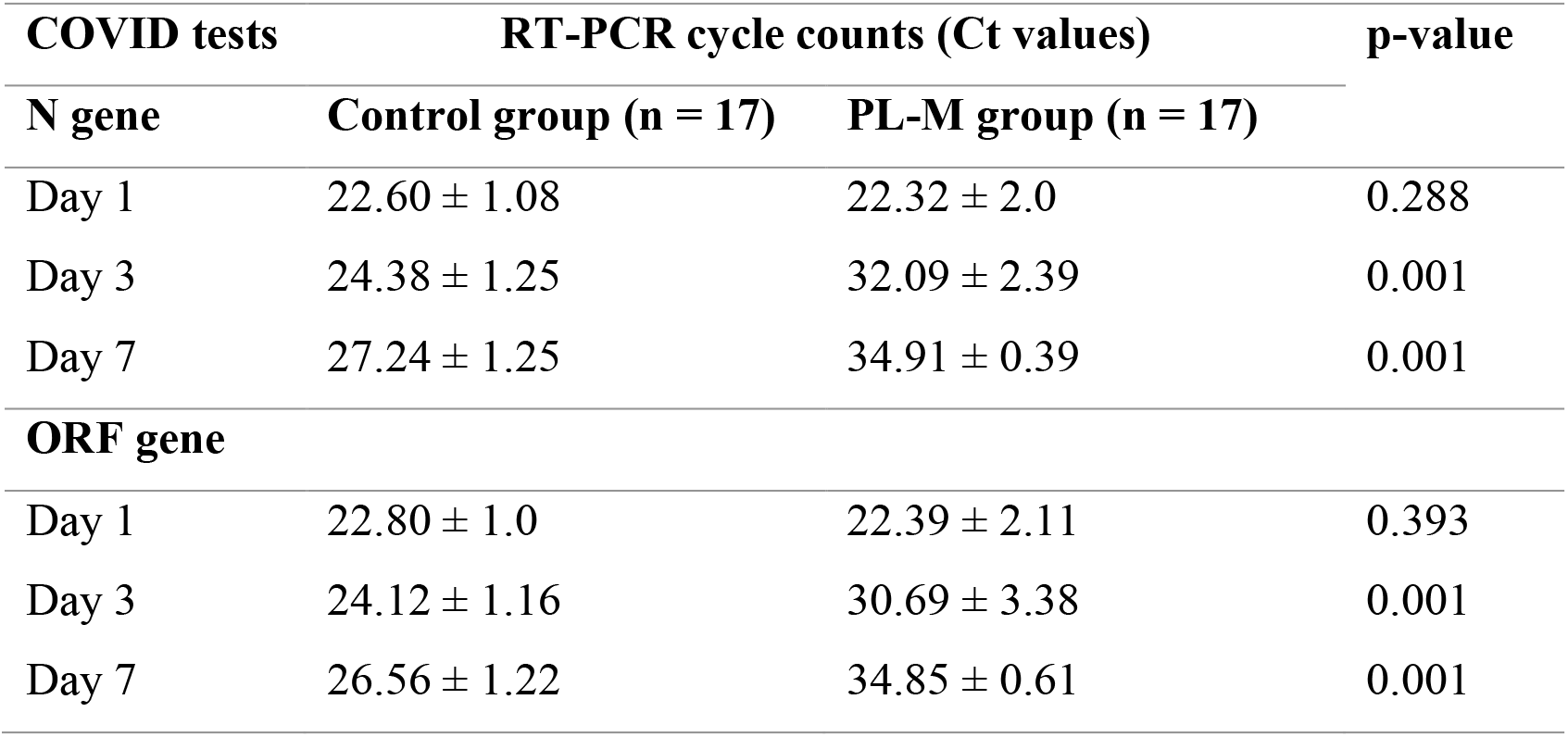
RT-PCR cycle counts for N and ORF gene at different visits

| COVID tests | RT-PCR cycle counts (Ct values) |  | p-value |
| --- | --- | --- | --- |
| N gene | Control group (n = 17) | PL-M group (n = 17) |  |
| Day 1 | 22.60 ± 1.08 | 22.32 ± 2.0 | 0.288 |
| Day 3 | 24.38 ± 1.25 | 32.09 ± 2.39 | 0.001 |
| Day 7 | 27.24 ± 1.25 | 34.91 ± 0.39 | 0.001 |
| ORF gene |  |  |  |
| Day 1 | 22.80 ± 1.0 | 22.39 ± 2.11 | 0.393 |
| Day 3 | 24.12 ± 1.16 | 30.69 ± 3.38 | 0.001 |
| Day 7 | 26.56 ± 1.22 | 34.85 ± 0.61 | 0.001 |

**Figure 1.**
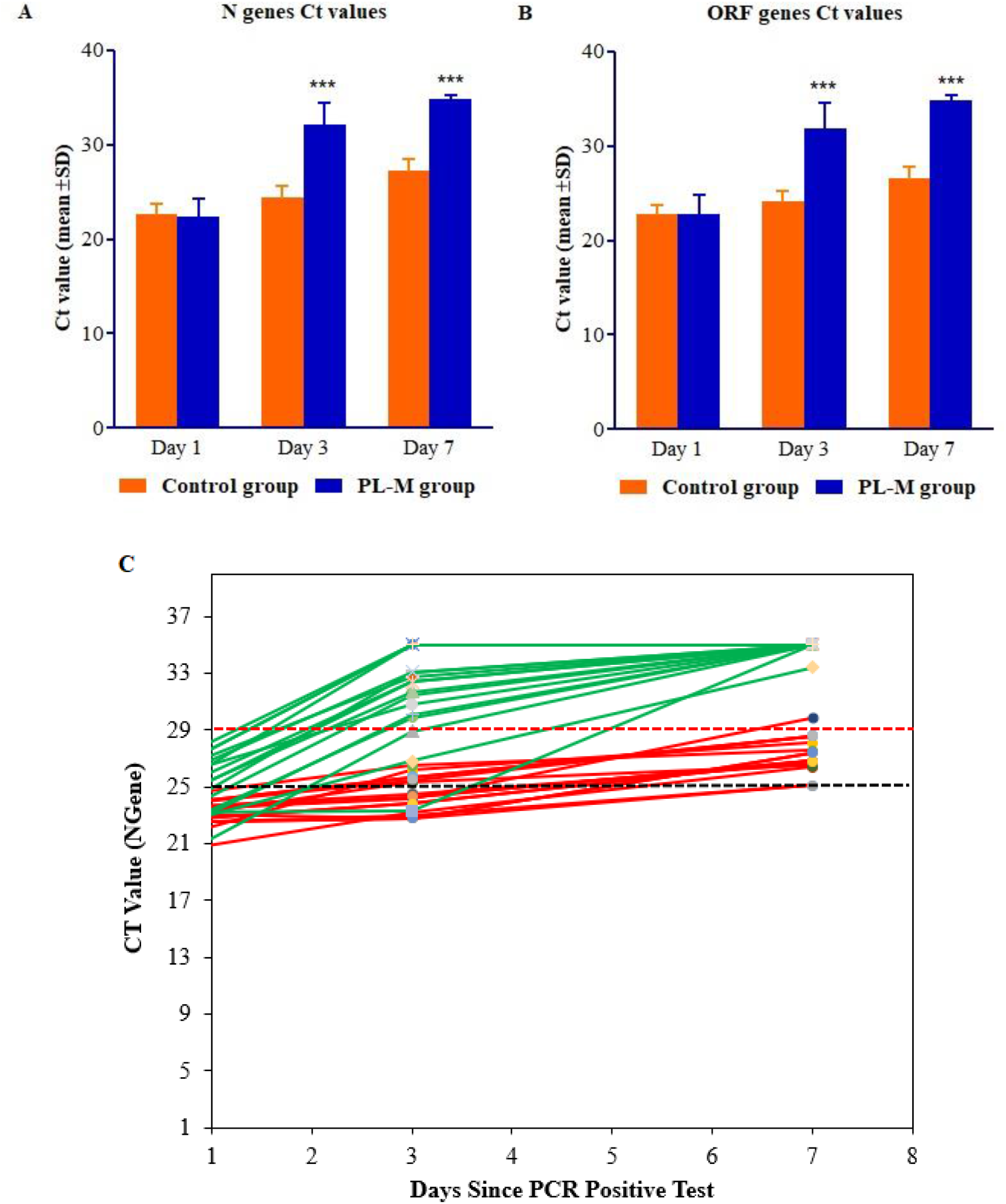
RT-PCR profiles of (**A**) the N gene, (**B**) the ORF gene, and (**C**) change in Ct value of individual subject in the placebo group (n=17) and PL-M group (n=17). RT-PCR profiles of N and ORF genes detected SARS-CoV-2 in COVID-19 patients on days 1, 3 and 7. On day 3, 82.35% of participants in the PL-M group had cycle counts for the N gene above the cut-off value of 29, while on day 7 all participants had cycle counts above the cut-off value. The majority of participants (94.12%) in the placebo group had cycle counts for the N gene below the cut-off value on day 7. Changes in RT-PCR cycle counts for N and ORF genes in the PL-M group at day 3 and 7 were statistically significant compared to the placebo group (p = 0.001).

### Assessment of safety

The changes in vital signs were assessed at all visits, and no abnormal changes in vital signs were observed in either study group. In both groups, all haematological and serum biochemical parameters showed no abnormal changes and were within the normal range. During the study period, no serious adverse events were reported.

### Binding of PL-M to Gal-3 using HSQC NMR

Having established the *in vitro* and *in vivo* effectiveness of PL-M against SARS-CoV-2 infectivity [23, 24], we sought insight on the molecular level. We hypothesized that PL-M functions *in situ* by binding to and antagonizing Gal-3, which is known to interact with SARS-CoV-2 to promote viral entry into cells [24]. To validate this proposal, we used NMR spectroscopy to assess binding interactions between PL-M and Gal-3, as well as between Gal-3 and SARS-CoV-2 S1 spike protein.

HSQC NMR spectra of ^15^N-labeled Gal-3 (^15^N-Gal-3) were measured as a function of PL-M concentration (0.3, 0.6, 1.2, 2.4 and 4.8 mg/mL). An ^15^N-^1^H HSQC spectral expansion is shown in Figure 2A for ^15^N-Gal-3 in the absence (peaks in black) and presence (peaks in red) of 1.2 mg/mL PL-M. During the titration, Gal-3 resonances were differentially chemically shifted and reduced in intensity (broadened), with some peaks becoming so broadened by the end of the titration that they cannot be observed. This observation alone demonstrates that Gal-3 binds to PL-M and indicates that the overall structure of Gal-3 is not significantly perturbed upon binding. Moreover, the fact that resonances are significantly broadened and minimally chemically shifted, PL-M binding to Gal-3 falls in the intermediate exchange regime on the chemical shift time scale, suggesting that the equilibrium dissociation constant (K_D_) lies in the 2 μM to 100 μM range [25]. The extent of resonance broadening is related to binding affinity and stoichiometry, in addition to any binding-induced changes in internal motions and conformational exchange. Therefore, one might look at this as binding avidity, or the net ability of PL-M to bind to Gal-3. Alternatively, or in addition, the apparent broadening could result from multiple binding modes given the heterogeneous nature of the PL-M polysaccharide. Either way, Gal-3 binds relatively strongly to PL-M.

**Figure 2.**
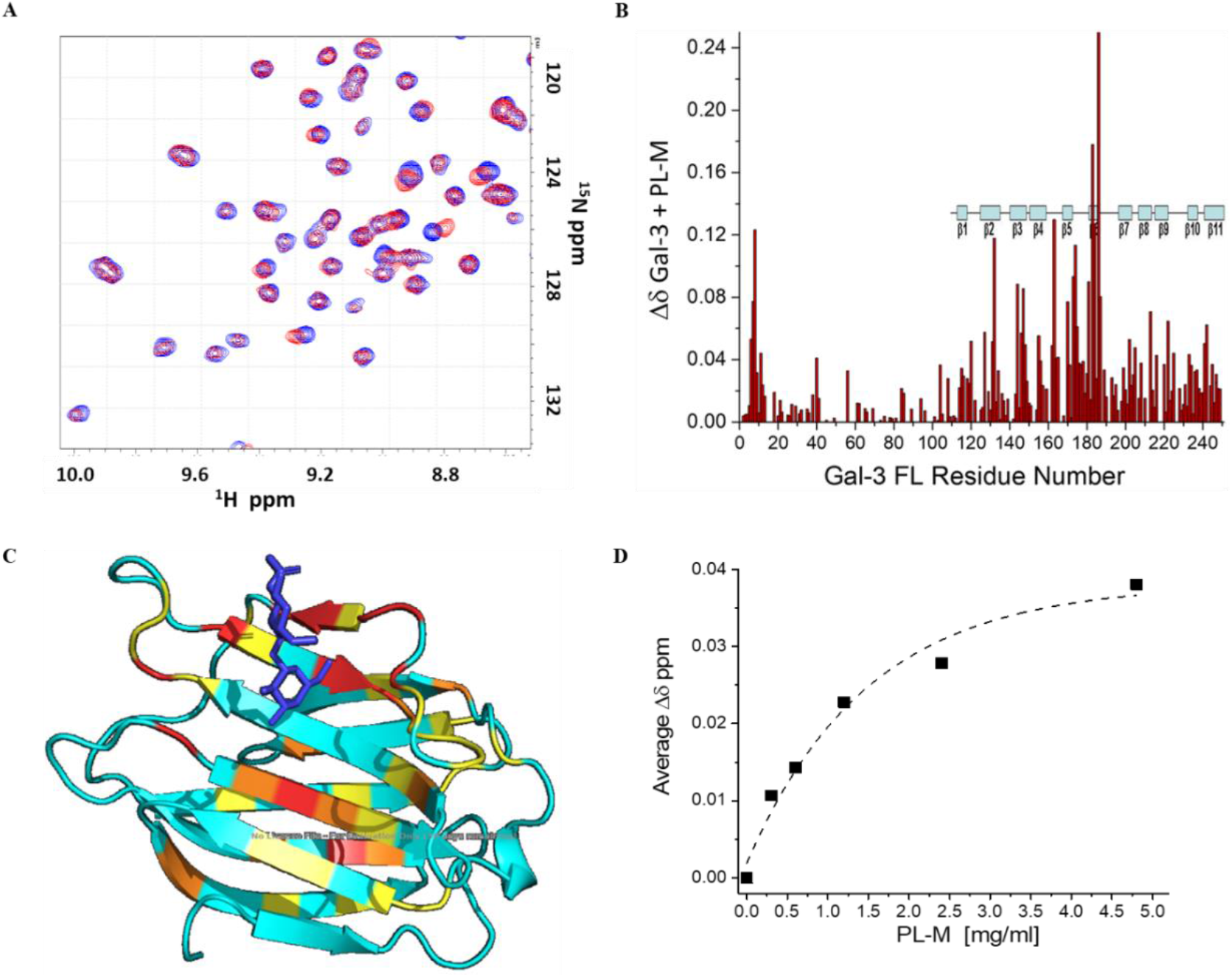
PL-M binding to Gal-3. (**A**) ^15^N HSQC expansions are overlaid for ^15^N-labeled Gal-3 (20 μM) in the absence (black peaks) and presence of 1.2 mg/ml PL-M (red peaks). (**B**) Chemical shift map (Δδ vs. the amino acid sequence of Gal-3) is shown for the binding of PL-M to Gal-3. Chemical shifts were internally referenced to DSS (4,4-dimethyl-4-silapentane-1-sulfonic acid), and chemical shift differences (Δδ) were calculated as [(Δ^1^H)^2^ + (0.25 Δ^15^N)^2^]^1/2^. Solution conditions were 20 mM potassium phosphate, pH 6.9. (**C**) The crystal structure of the Gal-3 CRD (pdb access code: 1A3K; Seetharaman et al., 1998) is shown with the largest Δδ values highlighted in red (> 2SD above the Δδ average), orange (between 1SD to 2SD above the Δδ average), yellow (between the average and 1SD above the Δδ average), and aqua (below the Δδ average). For orientation, a molecule of bound lactose is shown in dark blue in stick format. (**D**) Δδ values averaged over all Gal-3 residues are plotted vs the concentration of PL-M. Data were exponentially fitted as shown by the dashed line.

Even though PL-M binding-induced ^15^N-Gal-3 chemical shift changes, Δδ, are relatively small, they are useful to assess the binding epitope on the lectin. Figure 2B plots ^15^N-Gal-3 chemical shift changes, Δδ, vs. the amino acid sequence of Gal-3. The most shifted resonances arise from Gal-3 CRD residues in β-strands 3, 4, 5 and 6 that comprise the S-face β-sheet of the β-sandwich to which the β-galactoside lactose binds, as illustrated in Figure 2C, which shows the structure of the Gal-3 CRD (pdb access code: 1A3K) with the most shifted residues highlighted. This indicates that the PL-M binding epitope on Gal-3 is within the canonical sugar binding domain on the S-face of the CRD. Chemical shift changes within the N-terminal tail (NT) (residues 1-111) most likely result from PL-M binding-induced allosteric effects on the CRD F-face and modulation of interaction dynamics between the NT and CRD F-face [26]. Figure 2D plots chemical shift changes averaged over all Gal-3 resonances vs. the concentration of PL-M with the 50% saturation point in the plot falling at ∼1 mg/mL. However, because Gal-3 binding falls in the intermediate exchange regime, one cannot accurately determine binding affinity/avidity (or stoichiometry), other than the equilibrium dissociation constant, K_d_, falling in the ∼2 μM to ∼100 μM range [25]. Essentially the same binding results were observed with PL-M and ^15^N-labeled truncated Gal-3 CRD that is devoid of its N-terminal tail (data not shown).

### Binding of Gal-3 to SARS-CoV-2 S1 Spike Protein using HSQC NMR

To further validate our hypothesis, we used NMR spectroscopy to assess interactions between the viral spike protein S1 and Gal-3, both to the full-length lectin and to its truncated CRD form. HSQC NMR spectra of ^15^N-labeled full length Gal-3 (Gal-3 FL) were measured as a function of spike protein concentration (0.2, 0.4, 1, 2, 4 and 10 µM). ^15^N-^1^H HSQC spectra are shown in Figure 3A for ^15^N-Gal-3 FL in the absence (peaks in black) and presence (peaks in red) of 1 µM spike protein. As with PL-M binding, Gal-3 resonances are differentially chemically shifted and reduced in intensity (broadened), with some peaks becoming so broad by the end of the titration that they could not be observed, demonstrating that Gal-3 FL binding to the spike protein occurs in the intermediate exchange regime on the chemical shift time scale with an apparent K_D_ lying in the 2 μM to 100 μM range. Figure 3B shows that Gal-3 FL resonance broadening is about 50% at 1 µM spike protein, with effects within the NT and CRD occurring to the same extent. This suggests that the NT itself, and not only the CRD, may play a role in binding to the spike protein.

**Figure 3.**
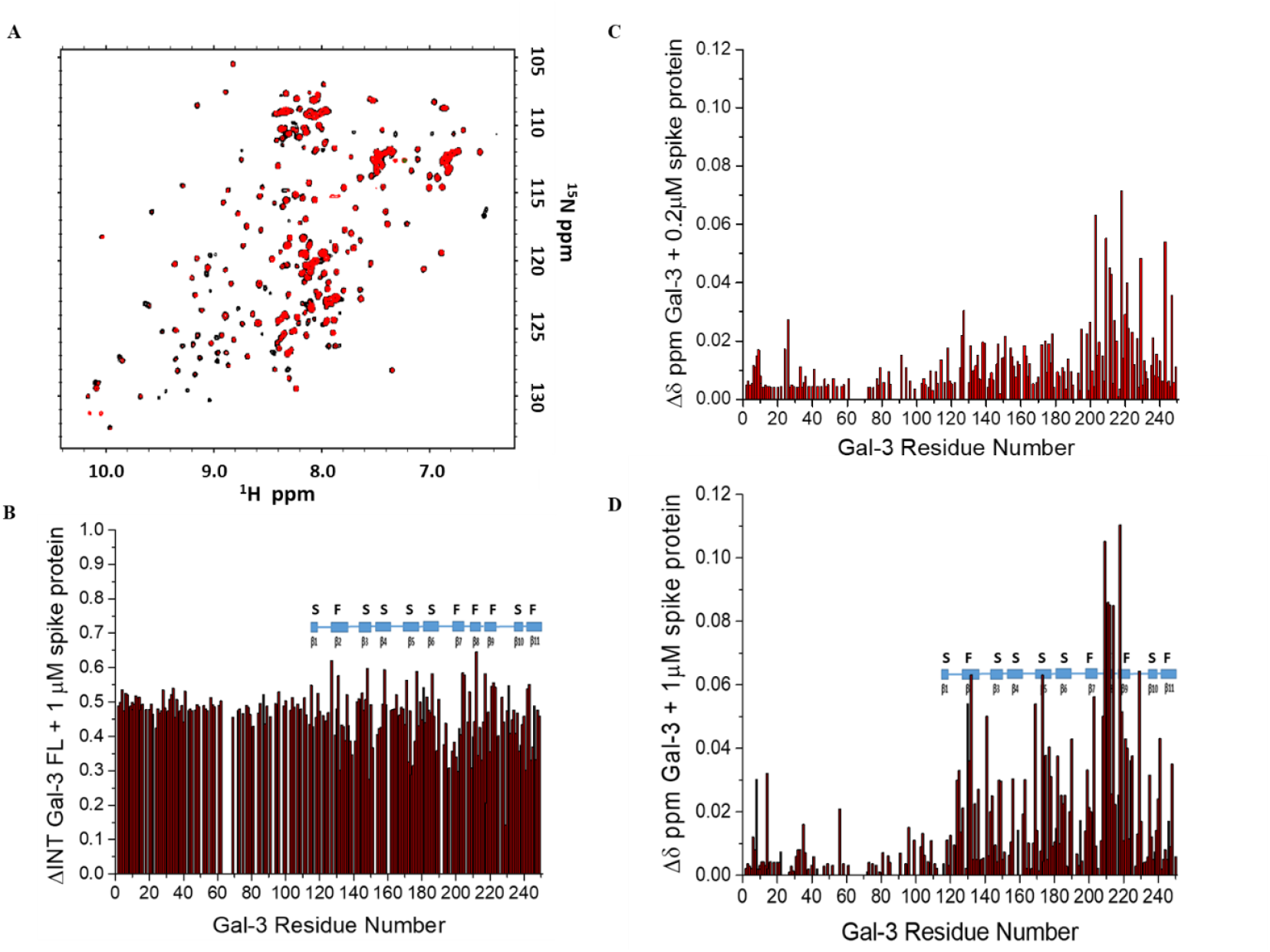
Gal-3 binding to SARS-CoV-2 spike protein. (**A**) ^15^N HSQC spectra are overlaid for ^15^N-labeled Gal-3 (20 μM) in the absence (black peaks) and presence of 1 µM S1 spike protein (red peaks). (**B**) ΔINT values that reflect resonance broadening due to the presence of the spike protein are shown vs. the Gal-3 FL residue number. (**C**) Δδ values calculated in the absence and presence of 0.2 µM spike protein are plotted vs. the Gal-3 residue number. (**D**) Δδ values calculated in the absence and presence of 1 µM spike protein are plotted vs. the Gal-3 residue number. Solution conditions were 20 mM potassium phosphate, pH 6.9.

Figure 3C plots ^15^N-Gal-3 FL chemical shift changes, Δδ, vs. the amino acid sequence of Gal-3 at the spike protein concentration of 0.2 µM. Here, it is important to note that while canonical sugar binding S-face residues are chemically shifted, the most shifted resonances actually arise from Gal-3 residues within β-strands 2, 7, 8 and 9 that comprise the F-face β-sheet to which the NT transiently interacts [26]. This indicates that the initial interaction site with the spike protein is primarily on the F-face. However, at 1 µM spike protein (Figure 3D), binding to the Gal-3 FL CRD S-face becomes more enhanced as one might expect.

HSQC spectra of ^15^N-labeled truncated Gal-3 CRD were also measured as a function of spike protein concentration (0.2, 0.4, 1, 2, 4 and 10 µM). ^15^N-^1^H HSQC spectra are overlaid in Figure S1A for ^15^N-Gal-3 in the absence (peaks in black) and presence (peaks in red) of 1 µM spike protein. Even though it appears that binding of both Gal-3 FL and truncated Gal-3 CRD to the spike protein occur to similar extents, as reflected in changes in resonance broadening (Figure S1B,C), Δδ values for truncated Gal-3 CRD at 1 µM spike protein are relatively very small and more scattered throughout the CRD, both S- and F-faces (Figure S1D), compared to Gal-3 FL. This suggests that Gal-3 FL with its N-terminal tail (NT) provides the better binding epitope(s) for interactions with the spike protein.

Figure S1C plots ΔINT values averaged over all Gal-3 resonances for both Gal-3 FL and Gal-3 CRD vs. the concentration of the spike protein. In both cases, broadening plateaus out at ∼4 µM spike protein, suggesting that binding is saturated at this concentration. Nevertheless, it is difficult to say that this is indeed the case, because binding interactions occur in the intermediary exchange regime where K_D_ values cannot be accurately determined [25]. Nevertheless, if binding were to saturate at ∼4 µM spike protein with Gal-3 at 20 µM, binding stoichiometry would be ∼5 molecules of Gal-3 per 1 molecule of spike protein, and thus ∼5 Gal-3 binding sites on the Omicron variant spike protein, a stoichiometry that seems reasonable.

### Lactose competes with Gal-3 to SARS-CoV-2 Spike Protein

A reduction in resonance broadening is a good indicator of a reduction in ligand binding when binding occurs in the intermediary exchange regime on the chemical shift time scale. In the absence of lactose, Gal-3 FL resonances are highly broadened, consistent with binding of Gal-3 FL to the SARS-CoV-2 S1 spike protein (Figure S2A). At 4 µM spike protein, Gal-3 resonance broadening (ΔINT) is less for residues within the NT and more so within the CRD of Gal-3 FL. This is not unusual, because the NT is dynamic and transiently interacts with the F-face of the CRD (Ippel et al., 2016). ΔINT values averaged over all residues 1-250 is 0.911, whereas ΔINT values averaged over residues 1-113 (NT) and 114-250 (CRD) are 0.816 and 0.972, respectively.

Addition of 10 mM lactose greatly decreases Gal-3 FL resonance broadening induced by its equilibrium binding to the spike protein (Figure S2B). Here, ΔINT values averaged over all residues 1-250, residues 1-113 and residues 114-250 are 0.473, 0.341, and 0.558, respectively, indicating that lactose competes with glycans on the surface of the spike protein for binding to Gal-3 FL. However, it is important to note that the lactose-induced reduction in Gal-3 FL resonance broadening occurs differentially, with some residues showing minimal if any change and others showing a large change (Figure S2B). This differential effect suggests that the Gal-3 glycan-binding epitope on the spike protein involves carbohydrates other than, and in addition to, β-galactosides like lactose. In fact, Lenza et al. reported that Gal-3 can bind to various glycosides on the spike protein [27].

With truncated Gal-3 CRD, resonances are also highly broadened in the presence of the spike protein, consistent with binding to the SARS-CoV-2 spike protein (Figure S2C), with the ΔINT averaged over residues 114-250 being 0.898. In this instance, addition of 10 mM lactose greatly decreases Gal-3 CRD resonance broadening (mostly non-differentially) throughout the entire CRD (Figure S2D), with the ΔINT averaged over residues 114-250 being 0.232. Thus, lactose is more effective at competing off truncated Gal-3 CRD from the spike protein compared to Gal-3 FL. Moreover, it appears that the β-galactoside lactose alone competes well with glycans on the surface of the spike protein for binding to truncated Gal-3 CRD. In this regard, the glycan binding profile for truncated Gal-3 CRD is different from that for Gal-3 FL, and indicates that Gal-3 FL binds a somewhat different complement of glycans on the spike protein.

## 4. Discussion

Patients with COVID-19 can be treated with corticosteroids, a combination of different antiviral drugs, healing plasma, certain antibiotics, and supportive care [28]. However, they were not very successful in preventing disease progression [29]. Therefore, any potential pharmacological candidates that can alleviate the progression of the disease and increase resistance to COVID-19 appear promising. As previously mentioned, human Gal-3 is crucial to the entry of SARS-CoV-2, and its blockage may prevent the progression of the disease. In an early in vitro study, we showed that PL-M lowered viral load and increased its clearance from Vero cells by inhibiting the entry of SARS-CoV-2 into Vero cells following PL-M binding to the NTD of the Gal-3 molecule [24]. Furthermore, in a small clinical study of 10 COVID-19 patients, we found that PL-M treatment resulted in a quick reduction in viral load and increased viral clearance without eliciting any serious adverse effects [23]. In this randomised, placebo-controlled clinical trial (n = 34), we found that PL-M treatment significantly reduced the duration of viral clearance in COVID-19 patients, as evidenced by an increase in RT-PCR cycle counts for SARS-CoV-2 N and ORF genes (mean cycle counts > 29 on days 3 and 7, p = 0.001). After 7 days of PL-M treatment, all participants were found to have higher cycle counts for both genes, which were well above the target cycle cutoff value (target cycle value 29). The placebo group had lower RT-PCR cycle counts for both genes through the end of the study (mean cycle counts < 29 in 7). On day 7, RT-PCR cycle counts revealed that all participants in the PL-M group had cycle counts greater than 29, while 94.12% of participants in the placebo group had cycle counts less than 29. Additionally, no major adverse effects or abnormalities in vital signs, haematological, or biochemical parameters were identified after 7 days of PL-M administration. Our findings were consistent with those of earlier in vitro and clinical studies [23, 24].

In this regard, we used ^1^H–^15^N HSQC NMR spectroscopy to determine the spike protein blocking mechanism of PL-M. Our NMR structural studies indicate that PL-M binds specifically to Gal-3 in the µM range. In relation to Gal-3 inhibitors, it is plausible that our compound interacts with Gal-3 in the same way as the structurally similar NTD of the S1 subunit of SARS-CoV-2. Overall, inhibition of Gal-3, as well as blockade of the NTD of the S1 subunit and ACE2 receptors, may be prospective anti-SARS-CoV-2 actions of PL-M.

## 5. Conclusions

Our findings shed light on the efficacy of PL-M in preventing SARS-CoV-2 infection as well as the mechanism by which it prevents viral entry into cells. In this study, we established that PL-M reduces viral load and increases viral clearance in patients with mild to severe COVID-19. In addition to exploring the clinical efficacy of PL-M, we also conducted 1H-15N HSQC NMR investigations to determine the mechanism of PL-M in preventing SARS-CoV-2 infection. NMR studies showed that PL-M has a high affinity for Gal-3. Overall, PL-M inhibits SARS-CoV-2 entry into cells through Gal-3 inhibition. Due to its great efficacy and tolerability, PL-M has the potential to be used in the treatment and prevention of COVID-19. However, larger-scale trials are required to prove the therapeutic feasibility and effectiveness of PL-M against COVID-19 disease.

## Data Availability

All data supporting this study are provided in the main article, supplementary information files, and upon request to the corresponding author.

## Acknowledgements

The authors are thankful to Murli Krishna Pharma and Dr. Madhumohan Rao for their support during the study. The authors wish to thank all the participants who participated in the study.

## Institutional Review Board Statement

The study was conducted according to the guidelines of the Declaration of Helsinki, and approved by the Institutional Ethics Committee (IEC) of ESIS Medical College and Hospital, Sanath Nagar, Hyderabad, India (ESIC Registration No: ECR/1303/Inst/TG/2019; Protocol ID: SRSD2001/01/01 and approval date: 01 Feb 2022). As per regulatory standards, the study was registered with the Clinical Trials Registry-India (CTRI/2022/03/040757) on March 3, 2022.

## Informed Consent Statement

This study used a randomised, placebo-controlled approach with human volunteers to assess the efficacy of the study intervention. All patients provided signed informed consent prior to any study-related procedure.

