## Supplementary material for "Galectin approach to lower covid transmission - Drug Development for clinical use": Figure S1, Figure S2: Supplementary file.docx

**Supplementary Figures:**

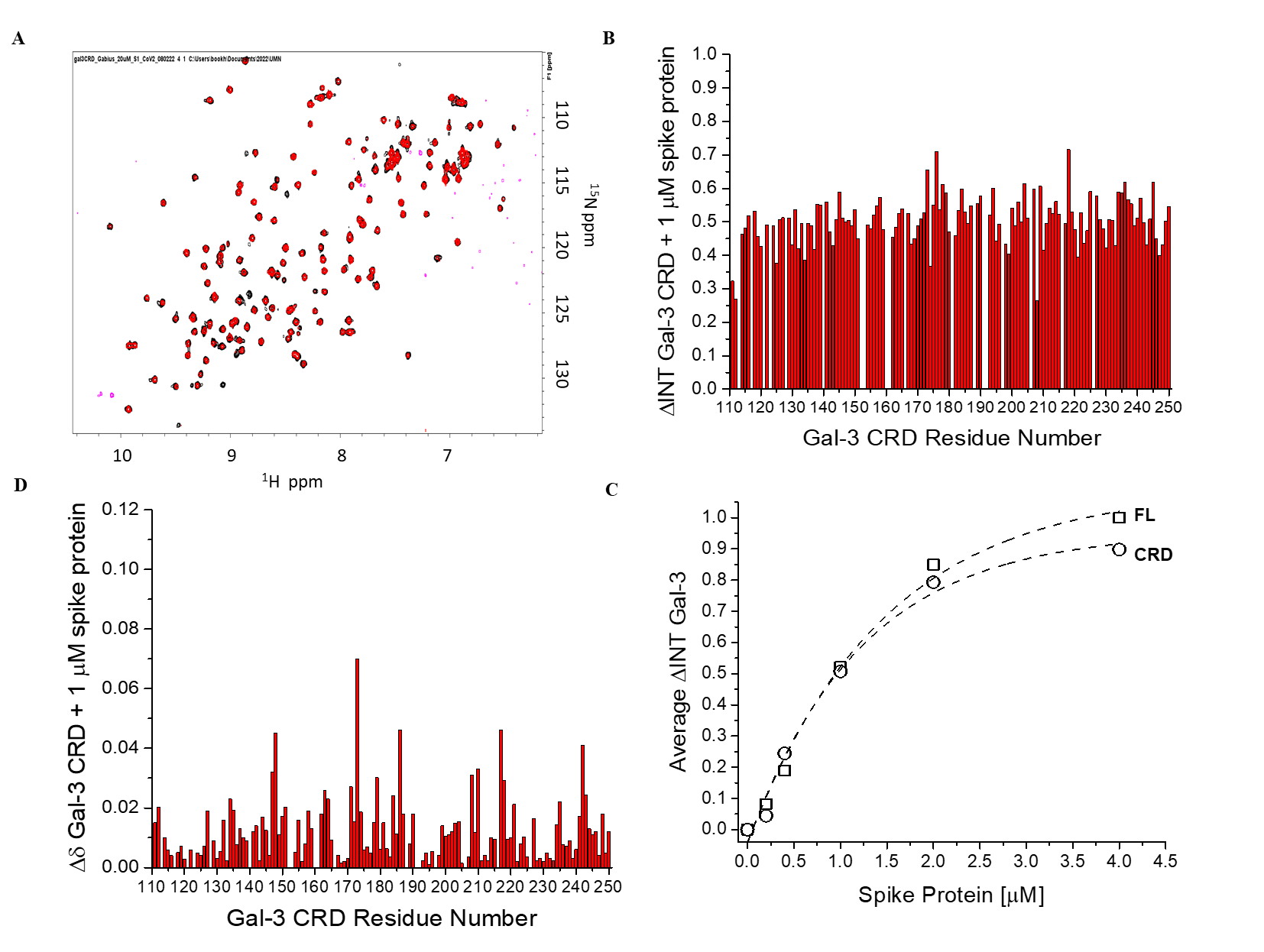

**Figure S1:** Interaction of Gal-3 to SARS-CoV-2 S1 Spike Protein using HSQC NMR spectroscopy

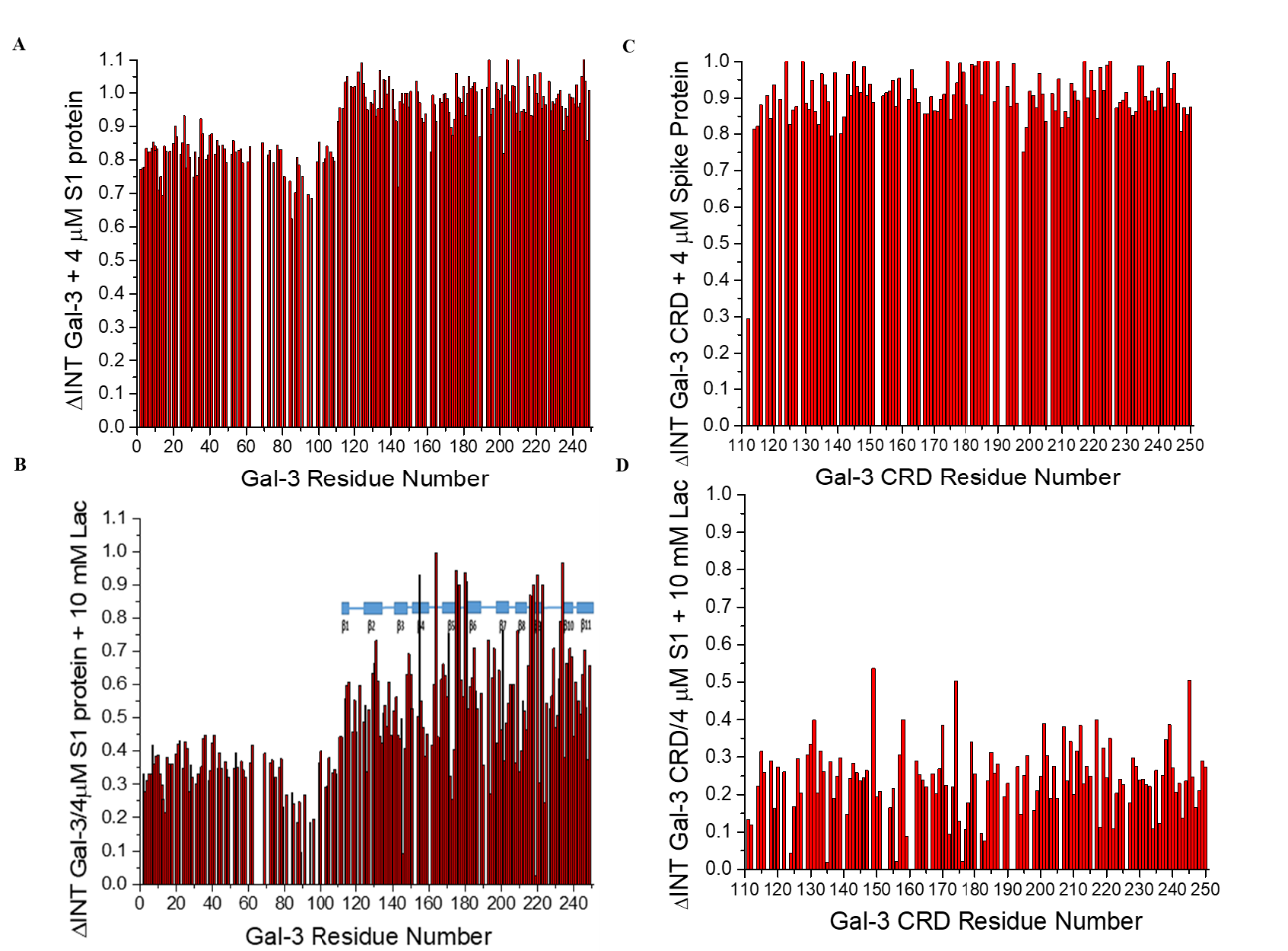

**Figure S2:** Lactose competes with Gal-3 to SARS-CoV-2 Spike Protein using HSQC NMR spectroscopy
